# Coinfection with *Strongyloides* and SARS-CoV-2: protocol for a systematic review

**DOI:** 10.1101/2023.01.30.23285219

**Authors:** Elena C Rosca, Carl Heneghan, Elizabeth A Spencer, Annette Plüddemann, Susanna Maltoni, Sara Gandini, Igho J Onakpoya, David H Evans, John M Conly, Tom Jefferson

## Abstract

**Rationale for the review:** COVID-19 treatment can worsen parasitic disease in patients with coinfection. Consequently, there is a need to investigate the infection with SARS-CoV-2 and *Strongyloides*. We aim to systematically review clinical and laboratory features of COVID-19 and *Strongyloides* coinfection, to investigate possible interventions and outcomes in this pathology. Also, we aim to identify difficulties in managing the parasitic disease manifestations in this context and to emphasize research gaps requiring further attention.

**Methods:** We will search two electronic databases – LitCOVID, and WHO COVID-19 and will include studies on SARS-CoV-2 and *Strongyloides* coinfection. We will adapt the WHO-UMC system for standardized case causality assessment to evaluate if using corticosteroids or other immunosuppressive drugs in COVID-19 patients determined acute strongyloidiasis manifestations.

**Expected results:** We will present the evidence in three distinct packages: study description, methodological quality assessment and data extracted. We will summarize the evidence and will draw conclusions as to the quality of the evidence.

## Introduction

Strongyloidiasis is a neglected disease caused by *Strongyloides stercoralis*, a soil-transmitted nematode (roundworm). It has been reported not only in tropical and subtropical regions but also in areas of lower endemicity in temperate climates.^1^An estimated 600 million people are infected worldwide,^2^ but precise data on prevalence are lacking in endemic countries. The unique characteristics of the *Strongyloides* are its ability to persist and replicate within a host for decades while producing minimal or no symptoms and its potential to cause life-threatening infections by dissemination and hyperinfection in the setting of immunosuppression that can become life-threatening.^3,4^

Strongyloidiasis presents a broad spectrum of clinical manifestations, with five main clinical pictures: (i) asymptomatic intestinal infection; (ii) acute infection with cutaneous manifestations and Loeffler’s syndrome; (iii) chronic intestinal disease with chronic anemia, eosinophilia, malabsorption, and chronic diarrhea; (iv) hyperinfection syndrome (HS); and (v) disseminated strongyloidiasis (DS).^5^

The majority of the infected individuals are asymptomatic, or they present with intermittent symptoms, mainly digestive complaints (from mild abdominal pain or diarrhea to more severe presentations mimicking inflammatory bowel disease), respiratory symptoms (cough, wheezing and asthma, chronic bronchitis), and dermatological sign (pruritus, rash). Also, patients may present systemic involvement with weight loss and cachexia.^4^ The immunocompromised individuals have an increased risk of developing HS and DS, which may be fatal.^4^ The most common trigger for HS is the use of corticosteroids,^6,7^ the syndrome being reported even after short courses of corticosteroids (e.g., six days)^8^, in low doses (e.g., 20 mg of prednisone/day).^9^ Other risk factors for HS and DS include immunosuppressive therapy, solid organ, and hematopoietic stem cell transplantation, HIV/AIDS, HTLV-1 infection, hematologic malignancies (e.g., leukemia, lymphoma), solid tumors, systemic collagen disease, chronic renal failure, and the use of H2 blockers and antacids.^5,10,11^

The Coronavirus Disease 2019 (COVID-19) has affected over 600 million people worldwide, with almost 6.5 million deaths.^12^ Therefore, a high number of patients with both, strongyloidiasis and SARS-CoV-2 is to be expected.

In patients with moderate and severe SARS-CoV-2 infection, the current therapies include administering anti-inflammatory agents with immunosuppressive effects like dexamethasone and tocilizumab.^13^ Therefore, in patients with coinfection, there is a high risk of developing HS and DS, ^13-16^, as immunosuppression can lead to worsening the parasitic disease.^13^

In this context, there is a need to investigate infection with SARS-CoV-2 and *Strongyloides*, as HS and DS are potentially severe conditions, with mortality rates of approximately 90% if left untreated.^17,18^ Furthermore, there are still knowledge gaps regarding many aspects of this coinfection.

Our objective was to systematically review the clinical and paraclinical features of the COVID-19 and *Strongyloides* coinfection, to investigate possible interventions and the outcomes in this pathology. Also, we aimed to identify the difficulties in managing HS and DS in this context and to emphasize research gaps that require further attention.

## Methods

We will conduct searches in the following electronic databases: LitCOVID and the World Health Organization (WHO) COVID-19. As the databases are specific for COVID-19, we used the following terms: “Strongyloides,” “Strongyloidiasis,” “Anguillulose,” and “Anguillulosis”; a search string to identify articles on SARS-CoV-2 infection will not be necessary. Also, we will screen for additional studies the reference lists of relevant articles. We will not set any language restrictions.

All references will be stored, organized, and managed with bibliographic software (EndNote 20, Clarivate Analytics, PA, USA).

We will include studies reporting on patients with SARS-CoV-2 infection and strongyloidiasis without any age, gender, or region restrictions.

We aim to include case reports, case series, and prospective or retrospective observational studies and interventional studies. If available, besides primary studies, we also plan to include systematic reviews. Conference abstracts will be included only if the authors did not publish a full article on the study.

From the included studies, we will extract the following information: publication details (authors, year, country), study type, and patient data (age, gender, country of origin, immunological status, comorbidities and medication, clinical signs and symptoms, the time between SARS-CoV-2 infection and strongyloidiasis manifestations, the time between SARS-CoV-2 infection and strongyloidiasis diagnosis, paraclinical findings, postmortem investigations, treatment, COVID-19 severity, evolution and outcome of coinfection, and the assumed mechanism of strongyloidiasis manifestations.

One reviewer (ECR) will extract data from the included studies, and these will be independently checked by a second reviewer (EAS).

We will adapt the WHO-UMC system for standardized case causality assessment^19^ in order to evaluate if the use of corticosteroids in COVID-19 patients determined the acute manifestations of strongyloidiasis. Basically, we plan to make a combined assessment taking into account the clinical, paraclinical, and pharmacological aspects of the case history and the quality of the documentation of observation. The causality categories included certain, probable/likely, possible, unlikely, or unclear links between the use of corticosteroids and strongyloidiasis. One reviewer (ECR) will assess the causality in the included studies, and a second reviewer with experience in the diagnosis and treatment of strongyloidiasis will independently check the causality assessment (JC). Disagreements will be resolved by consensus, or a third reviewer if needed. We will report the results of individual patients where possible. We will consider meta-analyses if appropriate.

### Data synthesis and reporting

The outcome will consist of the reactivation of strongyloidiasis in the context of SARS-CoV-2 coinfection. We will provide a narrative summary of the epidemiological, clinical, and paraclinical data and report the outcomes, including quantitative estimates where feasible and relevant. Where possible, compatible datasets may be pooled for meta-analysis. We will also report research and policy implications, dependent on the findings.

## Data Availability

All data produced in the present work are contained in the manuscript

## Conflict of interest

TJ disclosure is available here: https://restoringtrials.org/competing-interests-tom-jefferson/ CJH holds grant funding from the NIHR, the NIHR School for Primary Care Research, the NIHR BRC Oxford and the World Health Organization for a series of Living rapid reviews on the modes of transmission of SARs CoV 2, reference WHO registration No2020/1077093, and to carry out a scoping review of systematic reviews of interventions to improve vaccination uptake, reference WHO Registration 2021/1138353-0. He has received financial remuneration from an asbestos case and given legal advice on mesh and hormone pregnancy tests cases. He has received expenses and fees for his media work, including occasional payments from BBC Radio 4 Inside Health and The Spectator. He receives expenses for teaching EBM and is also paid for his GP work in NHS out of hours (contract Oxford Health NHS Foundation Trust). He has also received income from the publication of a series of toolkit books and appraising treatment recommendations in non-NHS settings. He is the Director of CEBM, an NIHR Senior Investigator and an advisor to Collateral Global.

DHE holds grant funding from the Canadian Institutes for Health Research and Li Ka Shing Institute of Virology relating to the development of Covid 19 vaccines and the Canadian Natural Science and Engineering Research Council concerning Covid 19 aerosol transmission. He is a recipient of World Health Organization and Province of Alberta funding which supports the provision of BSL3 based SARS CoV 2 culture services to regional investigators. He also holds public and private sector contract funding relating to the development of poxvirus based Covid 19 vaccines, SARS CoV 2 inactivation technologies, and serum neutralization testing.

JMC holds grants from the Canadian Institutes for Health Research on acute and primary care preparedness for COVID 19 in Alberta, Canada and was the primary local Investigator for a Staphylococcus aureus vaccine study funded by Pfizer, for which all funding was provided only to the University of Calgary. He is a co-investigator on a WHO funded study using integrated human factors and ethnography approaches to identify and scale innovative IPC guidance implementation supports in primary care with a focus on low resource settings and using drone aerial systems to deliver medical supplies and PPE to remote First Nations communities during the COVID 19 pandemic. He also received support from the Centers for Disease Control and Prevention (CDC) to attend an Infection Control Think Tank Meeting. He is a member and Chair of the WHO Infection Prevention and Control Research and Development Expert Group for COVID 19 and the WHO Health Emergencies Programme (WHE) Ad hoc COVID 19 IPC Guidance Development Group, both of which provide multidisciplinary advice to the WHO, for which no funding is received and from which no funding recommendations are made for any WHO contracts or grants. He is also a member of the Cochrane Acute Respiratory Infections Group.

SM is a pharmacist working for the Italian National Health System since 2002 and a member of one of the three Institutional Review Boards of Emilia-Romagna Region (Comitato Etico Area Vasta Emilia Centro) since 2018.

AP holds grant funding from the NIHR School for Primary Care Research. IJO, EAS and ECR have no interests to disclose.

SG is an epidemiologist/biostatistician group leader at the European Institute of oncology in Milan (Italy) and holding a grant funded by the European Union’s Horizon Europe Research and Innovation Programme under Grant Agreement No 101046016 (Eucare project: EUROPEAN COHORTS OF PATIENTS AND SCHOOLS TO ADVANCE RESPONSE TO EPIDEMICS).

